# Muscle activation assessment using ultrasound time-harmonic elastography and tonic vibration reflex*

**DOI:** 10.1101/2025.09.09.25335388

**Authors:** Hossein S. Aghamiry, Tom Meyer, Stefan Klemmer Chandía, Pascal Engl, Giacomo Valli, Yanglei Wu, Eduard Kurz, René Schwesig, Thomas Bartels, Heiko Tzschätzsch, Jing Guo, Ingolf Sack

**Affiliations:** Department of Radiology, Charité – Universitatsmedizin Berlin, 10117, Berlin, Germany; Department of Engineering and Natural Sciences, University of Applied Sciences Merseburg, 06217 Merseburg, Germany; Department of Clinical and Experimental Sciences, University of Brescia, 25121, Brescia, Italy; Department of Orthopaedic and Trauma Surgery, Martin-Luther-University Halle-Wittenberg, 06120, Halle (Saale), Germany; MVZ Sportklinik Halle GmbH, 06108 Halle (Saale), Germany; Institute of Medical Informatics, Charité – Universitätsmedizin Berlin, 10117, Berlin, Germany

**Author notes:** Corresponding author; (Hossein S. Aghamiry). Funding by AIF FKZ KK5611902 BM4 (MUSKEL), German Research Foundation (DFG) projects 513752256 FOR5628, CRC1340, GRK2260 BIOQIC, and GU 172614-1 is greatly appreciated. These sponsors had no role in the study design, collection, analysis and interpretation of data, writing of this manuscript or the decision to submit the article for publication. Email address (Tom Meyer), (Stefan Klemmer Chandía), (Pascal Engl), (Giacomo Valli), (Yanglei Wu), (Eduard Kurz), (René Schwesig), (Thomas Bartels), (Heiko Tzschätzsch), (Jing Guo), (Ingolf Sack).

**Keywords:** Ultrasound elastography, time-harmonic elastography, tonic vibration reflex, muscle stiffness, vastus lateralis, shear wave speed, muscle function

## Abstract

**Background:** Muscle activation is associated with increased tissue stiffness as measured by elastography in diagnostic applications. For this reason, we present ultrasound time-harmonic elastography (THE) applied to the vastus lateralis (VL) muscle during passive tonic vibration reflex (TVR) and active voluntary contraction (VC) stimulation to test whether TVR can serve as a stimulation method for functional assessment of skeletal muscle stiffness.

**Methods:** Twenty-five asymptomatic volunteers (8 females, mean age 34 ± 8 years) underwent five consecutive THE examinations of the VL at three VC loads (15, 22, and 37 N) and during TVR stimulation with 100 Hz frequency and 200 µm (low) and 400 µm (moderate) amplitude. Using standard line-by-line ultrasound, THE acquired the induced shear waves of 60, 70, and 80 Hz frequency with a frame rate of 100 Hz. Shear wave speed (SWS) was reconstructed as a proxy for muscle stiffness and statistically analyzed with repeated-measures ANOVA and nonparametric Friedman tests.

**Results:** SWS increased significantly from 1.66±0.11□m/s at rest to 1.79±0.12□m/s, 1.93±0.13 m/s, and 2.16±0.12 m/s with 15, 22 and 37 N VC load (*p*< 10^-3^). Similar effects were observed during TVR activation with increases to 1.93±0.13 m/s and 2.18±0.14 m/s for low and moderate TVR amplitudes (*p* < 10^-3^). Increase of SWS at moderate TVR amplitudes correlated with that of 37 N VC load (*r* = 0.67, *p* < 10^-3^).

TVR-induced stiffness changes at 100 Hz vibration frequency and moderate amplitude can substitute the more subjective VC forces for muscle function testing. TVR stimulation combined with skeletal-muscle THE may be a useful tool for the routine clinical assessment of stiffness during muscle activation.

## 1. Introduction

Muscle activation is, by definition, associated with muscle fiber contraction and increased tissue stiffness (Fung, 2013). Tissue stiffness can be measured non-invasively by elastography based on MRI (Sack, 2023) or medical ultrasound (Tzschätzsch et al., 2024). Ultrasound-based elastography provides instantaneous feedback in real-time which is particularly beneficial for muscle function tests (Catheline et al., 2004; Deffieux et al., 2008; Gennisson et al., 2010; Parker et al., 2010; Eby et al., 2013; Doyley and Parker, 2014). Acoustic radiation force impulse (ARFI) elastography shows potential for capturing skeletal muscle anisotropy during activation (Hall et al., 2013; Song et al., 2013) while time-harmonic elastography (THE) can map stiffness across muscle groups in a single scan (Yang et al., 2024). THE uses continuous harmonic vibrations (20-80 Hz) to induce propagating shear waves. By encoding vibrations at multiple frequencies using a conventional line-by-line ultrasound scanner and stroboscopic sampling to handle aliased oscillations, THE generates high-resolution shear wave speed (SWS) maps reflecting tissue stiffness (Meyer et al., 2024). This approach uses conventional line-by-line ultrasound equipment, offering a cost-effective, versatile solution for full-field stiffness assessment.

Tonic vibration reflex (TVR) leads to involuntary muscle contractions due to sustained mechanical vibrations. Mediated by repetitive activation of muscle spindle Ia afferents, TVR enhances proprioception, facilitates motor control, and modulates muscle tone, offering potential for improving neuromuscular function and rehabilitation outcomes (Eklund & Hagbarth, 1966; Burke et al., 1972; Marín & Rhea, 2010). Previous research confirmed that TVR is influenced by vibration frequency, intensity, and neuromuscular excitability (Martin & Park, 1997). Lower frequencies (approximately 15–35 Hz) were shown to induce muscle activation, especially during whole-body vibration (WBV) (Zaidell et al., 2013). WBV studies suggest that the frequency range of 30–50 Hz effectively maximizes reflexive activation of extremity muscles (Marín & Rhea, 2010) while higher frequencies (approximately 50–150 Hz) elicit a strong TVR when applied to tendons or the muscle belly (Eklund & Hagbarth, 1966; Poenaru et al., 2016). Focal muscle vibrations in the tens of hertz regime can directly stimulate muscle fibers via propagating mechanical waves (Guang et al., 2018). In general, the optimal frequency to elicit TVR appears to be muscle-specific and depends on the stimulation method, frequency, amplitude, and muscle size (Poenaru et al., 2016).

Unlike TVR, active muscle contraction and co-contraction such as voluntary contraction (VC) in muscle function tests involve active force generation against external loads (Brandenburg et al., 2014). In this manner, VC forces can be generated directly or indirectly by the agonist or antagonist muscle, respectively. By using co-contraction of agonist and antagonist muscles at a joint movement is restricted and stiffness is increased. However, VC is subjective and induces short-term force fluctuations, limiting the consistency of muscle function tests in elastography (Yang et al., 2024). In contrast, although TVR provides a more standardized stimulation protocol, its physiological effects remain unclear. For example, the actual level of induced muscle activation is uncertain and may vary between individuals depending on factors such as limb size and tissue composition.

The vastus lateralis (VL) muscle, part of the quadriceps, is a key element of the upper leg’s motor unit and ideal for muscle function tests due to its superficial location and important role in knee extension (Siracusa et al., 2019). In addition, its easy accessibility and relatively uniform fiber alignment made it a practical and suitable target for our tests. VL stiffness correlates with both muscle functionality status (fatigue, strength) and functional changes during contraction (Xu et al., 2018; Tang et al., 2020). To advance muscle function testing, this study introduces TVR as a basic stimulation paradigm for elastography of skeletal muscle. While the role of elastography-inherent mechanical vibrations in stimulating TVR remains largely unexplored, the harmonic vibrations used by THE are considered to be below the thresholds of amplitudes and frequencies that significantly induce TVR. To induce TVR, a 100-Hz harmonic vibration tone was focally applied to the patellar tendon with amplitudes sufficiently high to stimulate the muscle activation. To prevent any interference with SWS measurement, the scanner frame rate was synchronized with the TVR vibration.

The aim of this study was to evaluate TVR as a muscle activation paradigm in elastography using THE in VL. Comparison of stiffness changes during TVR with established VC may provide an easy-to-use and objective method for assessing skeletal muscle function in the clinical context. We hypothesized that TVR can consistently elicit changes in muscle stiffness similar to subjective voluntary loads, demonstrating the utility of reflex-based stimulation for muscle function testing.

## 2. Materials and Methods

### 2.1. Subjects

25 asymptomatic volunteers (17 males, 8 females; mean age: 34 ± 8 years; Body Mass Index (BMI): 23.4 ± 2.9 kg/m^2^) without known musculoskeletal injury or neuromuscular disease were studied. Also, volunteers were asked to confirm that they had not engaged in any strong physical activity within 24 hours prior to the measurements. In 20 participants the dominant leg was studied. The study was approved by our institutional review board (EA-number: EA4/040/22) and all participants provided informed written consent before the study.

### 2.2. Experimental Setup and Study Protocol

Participants underwent THE in the right VL under six conditions: (1) rest (passive), (2–4) three VC levels and (5–6) two TVR stimulation levels. Each measurement of 40 s was separated by 2 min rest to minimize fatigue or adaptation. Subjects were seated upright with the knee fixed at 90° flexion (Figure 1a). An elastic strap secured the ultrasound probe over the mid-thigh to ensure consistent contact and orientation, while the THE-actuator—a custom 3D-printed half-cylindrical structure connected to a standard bass shaker (Sinustec ST-BS 250)—was positioned beneath the thigh (Figure 1a & 1b). THE measurements were performed at 70% of the distance between the proximal and distal myotendinous junctions, corresponding to a location closer to the distal end. For VC, three external loads were applied at the ankle, with force values of 15, 22, and 37 N. These values were determined through precise measurement using a force gauge (Pesola 80049) at the rope attached to the ankle, accounting for friction and other damping effects. These loads were selected based on a percentage of the load that, during the setup design phase, produced SWS values approximately matching those observed during TVR in one of the volunteers. Participants were instructed to resist any load-induced knee flexion during the scan. TVR-THE was performed under resting conditions while additionally applying a 100-Hz continuous tone using a normal exciter speaker (Sinustec St-Bs 250 bass-shaker) to the patellar tendon to induce involuntary VL contraction. 100 Hz with 200 and 400 µm peak-to-peak displacement amplitude (predefined and measured using a laser interferometer, Sick OD 5000 C30W05) was chosen to elicit a strong TVR stimulation. The TVR frequency was matched with the ultrasound frame rate in order to prevent any motion interference with radio frequency (RF) data acquisition.

**Figure 1:**
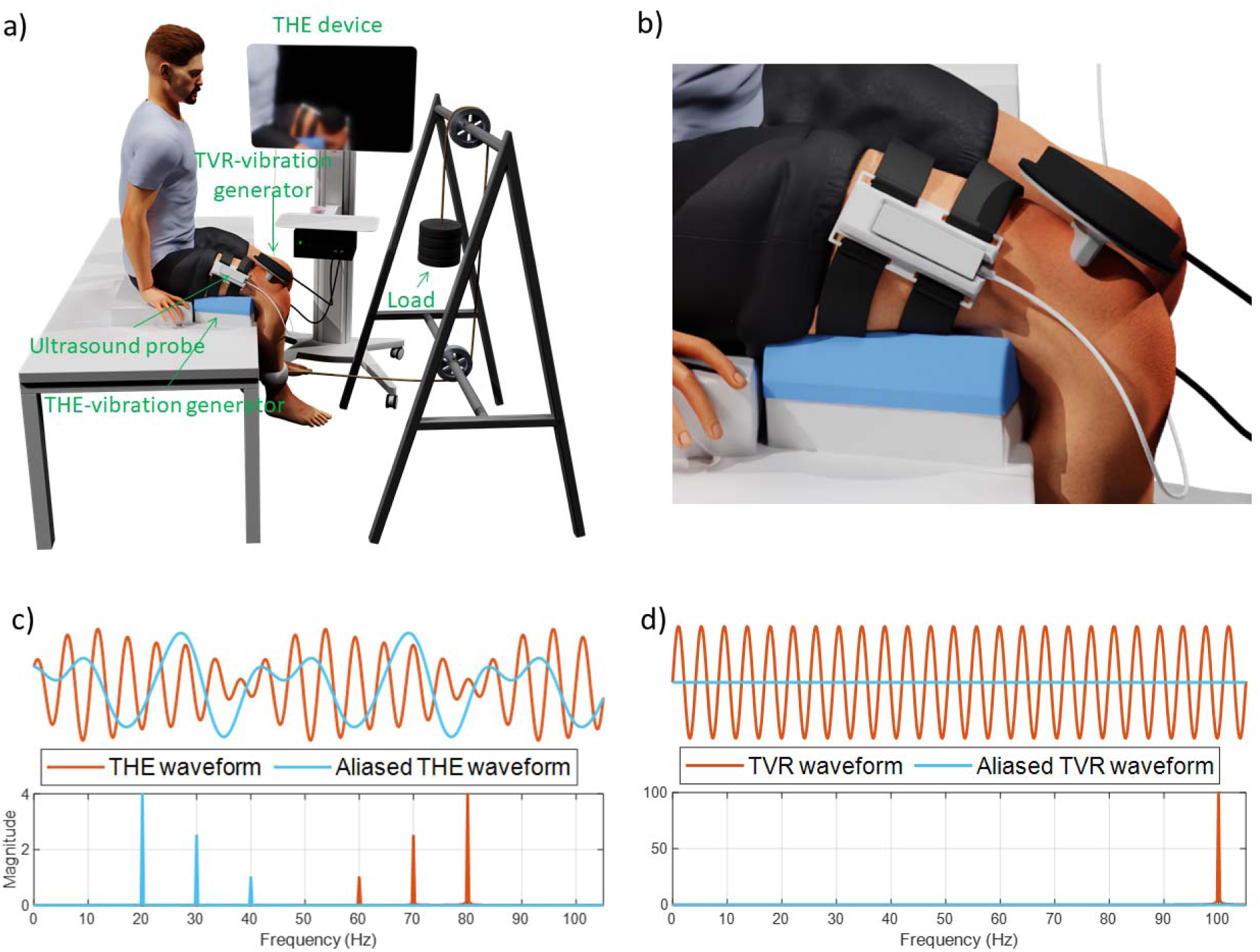
Experimental setup. (a) THE vibration generator beneath the thigh generates shear waves in the whole thigh. An elastic belt secures the ultrasound probe for consistent positioning. For VC trials, participants resist 15–37 N weights attached above the ankle while seated. For TVR trials, no external weight (0 N) was used; a 100 Hz vibrator (200 µm or 400 µm amplitude) at tendon evoked involuntary contraction. Note that, the TVR vibration generator was held by the operator to enhance measurement repeatability and ensure the subject remained in an upright seated position. (b) Close-up of the experimental setup. (c) THE harmonic waveform and frequency spectrum. Note that the three superimposed THE-vibration frequencies 60, 70 and 80 Hz are aliased to 40, 30 and 20 Hz due to the low ultrasonic sampling rate of 100 Hz. This aliasing is corrected during the post-processing. (d) TVR harmonic waveform and frequency spectrum.

### 2.3. Time-Harmonic Elastography (THE)

The vibration generator beneath the thigh provided continuous harmonic excitations of superimposed frequencies of 60, 70, and 80 Hz (Figure 1c). Shear waves were recorded by a clinical ultrasound system (GAMPT, Merseburg, Germany) and a linear probe (LF115H60A3; 5– 11 MHz; 54.6 mm field of view). The system operated in a line-by-line mode at 100 frames/s with 82 scan lines/frame. Data were recorded over 40 s; each 1 s segment (100 frames) was processed to generate one averaged SWS map yielding 40 SWS maps that depicted stiffness changes over time. Mean SWS values were extracted from a region of interest (ROI), and quintiles (Q10, Q50, Q90) were calculated per condition using all 40 maps. For TVR measurements, 8s of a stable plateau in mean SWS were used because reliable reflex measurement requires a pre-activation period.

ROIs were manually defined within homogeneous VL regions, avoiding muscle boundaries to minimize edge effects. To ensure signal quality, SWS values at spatial points with a z-score below 3 were excluded from the final map. The z-score, indicating sharpness of the spectral peaks, was computed from the temporal Fourier spectrum at each location across THE frequencies and then averaged. This filtering removed low-quality pixels from the final SWS time-average.

### 2.4. Data Processing

As illustrated in Figure 2, the acquired RF data were converted to SWS maps as follows:

1. **Displacement estimation:** Axial displacements between consecutive RF frames (Figure 2a) were estimated using Kasai’s phase shift algorithm (Kasai et al., 1985) with 1.5 mm windows to detect subtle tissue movements from harmonic vibrations (Figure 2b). Truncated singular value decomposition (SVD) denoising was applied to the reshaped spatio-temporal data by retaining the eigenvalue images of ranks 2 to 20.
2. **Frequency realignment:** Since 100 Hz frame rate resulted in 50 Hz Nyquist frequency, 60, 70, and 80 Hz vibrations were undersampled and aliased to 40, 30, and 20 Hz, respectively. A controlled aliasing technique (Tzschatzsch et al., 2016b) leveraged the precise knowledge of vibration frequencies and sampling rate to computationally shift these aliased peaks back to their true 60, 70, and 80 Hz positions.
3. **Harmonic component isolation:** Each harmonic component was then isolated using third-order Butterworth bandpass filters with a ±1.5 Hz bandwidth, yielding complex, frequency-resolved wavefields per vibration frequency (Figure 2c).
4. **Directional filtering:** Wavefields were decomposed into four in-plane directions (up, down, left, right) using steerable directional filters (Manduca et al., 2003). This filtering separated shear waves, suppressed compression artifacts using a Butterworth band-pass filter (10–200□Hz; 2nd-order high-pass, 3rd-order low-pass), and mitigated oblique incidence effects. Only the vertical components were used for SWS recovery, aligning with the direction of motion estimation and primary shear wave propagation relevant to VL muscle stiffness (Figure 2d).
5. **SWS reconstruction:** Local wavenumbers were calculated from the spatial gradient of the wrapped phase of the retained wavefield components. These were processed using amplitude-weighted multifrequency k-MDEV inversion (Tzschatzsch et al., 2016a) to generate time-resolved SWS maps (Figure 2e).

**Figure 2:**
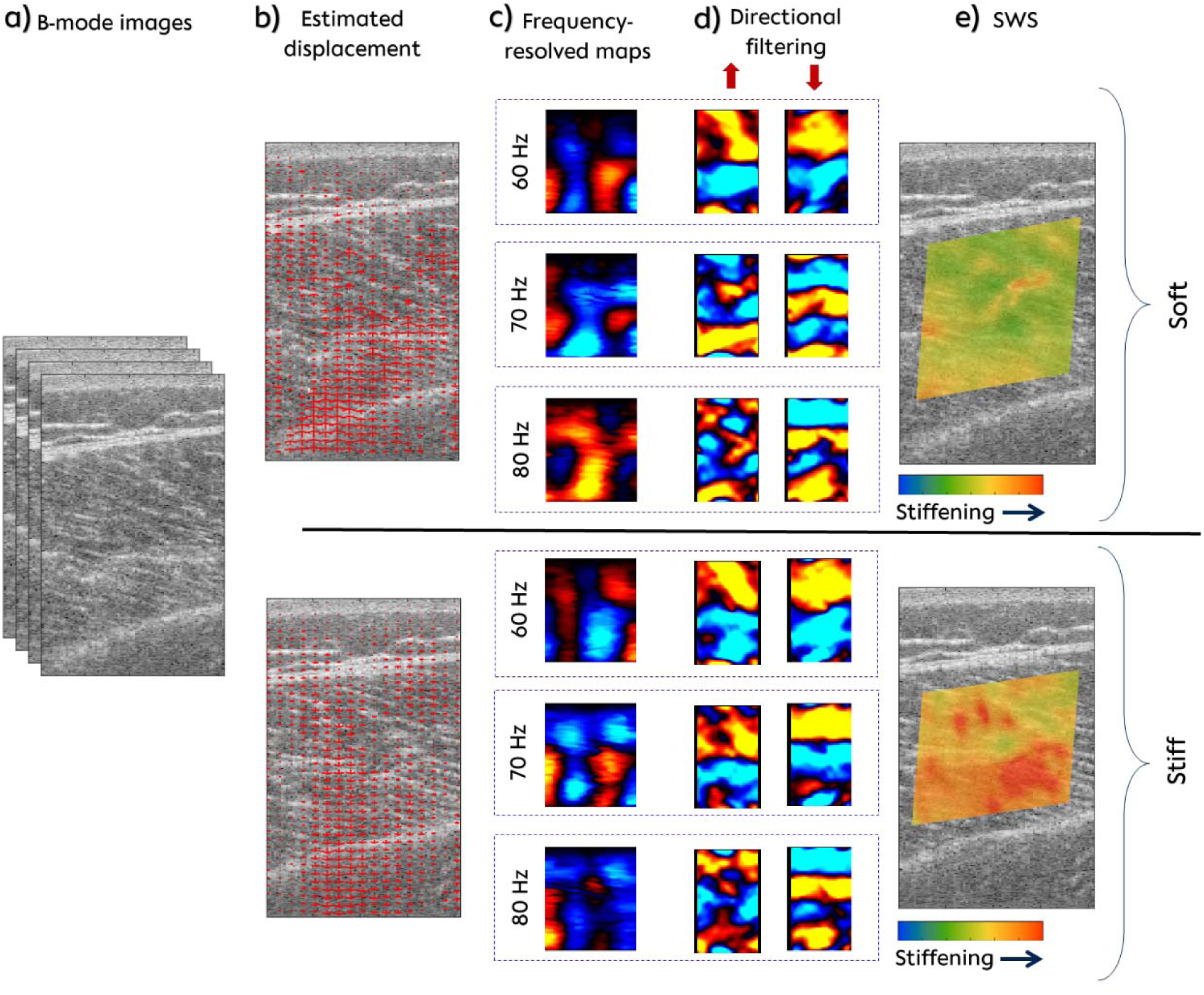
THE processing workflow for two conditions of muscle stiffness (indicated on the right-hand-side as Soft and Stiff). (a) B-mode ultrasound images of VL. (b) Axial tissue displacements (red arrows) from Kasai’s autocorrelation algorithm, overlaid on the B-mode. (c) SVD-denoised, realigned, alias-corrected, and bandpass-filtered signals yielded complex monochromatic wavefields at 60, 70 and 80 Hz frequency. (d) Directionally filtered isolated shear waves running into vertical directions (↑,↓). (e) Note, only vertically propagating wave components corresponding to the direction of displacement estimation were used as an input for multifrequency k-MDEV inversion to reconstruct SWS maps. Example SWS maps (colored patches) in a volunteer showing VL muscle softening (lower stiffness, top row) and stiffening (higher stiffness, bottom row) at baseline and during TVR, respectively.

### 2.5. Statistical Analysis

Statistical analyses involved both parametric and nonparametric tests to ensure robust results. Normality and sphericity were assessed using Shapiro–Wilk and Mauchly’s tests, respectively, applying Greenhouse–Geisser corrections when needed. Repeated-measures ANOVA and Friedman tests evaluated overall differences across conditions (Rest, VC at 15□N, 22□N, 37□N, TVR 200□µm, and TVR 400□µm). Post-hoc pairwise comparisons were performed using paired t-tests with Bonferroni corrections and Wilcoxon signed-rank tests as appropriate. Pearson correlation assessed relationships between VC and TVR stiffness at moderate and high intensities. Subgroup analyses by BMI, sex, age, and leg dominance used mixed-model ANOVA or nonparametric equivalents (Kruskal–Wallis and Mann–Whitney U tests). All tests were two-sided (α=0.05), with effect sizes reported as Cohen’s d (small: 0.2, medium: 0.5, large: 0.8, very large: 1.2, huge: 2.0) (Cohen, 1988; Sawilowsky, 2009).

## 3. Results

### 3.1. Time-Resolved SWS and B-mode Changes during TVR Stimulation

Figure 3 illustrates data from a representative volunteer during 400 µm TVR stimulation, detailing temporal changes in muscle SWS and B-mode ultrasound appearance. Figure 3a shows the average SWS over time within a selected ROI (outlined with dashed lines in panels e–g), with the shaded area indicating the standard deviation. SWS initially remained stable, then increased sharply around 10–15 s post-TVR onset, indicating the latency of reflex-induced stiffening.

**Figure 3:**
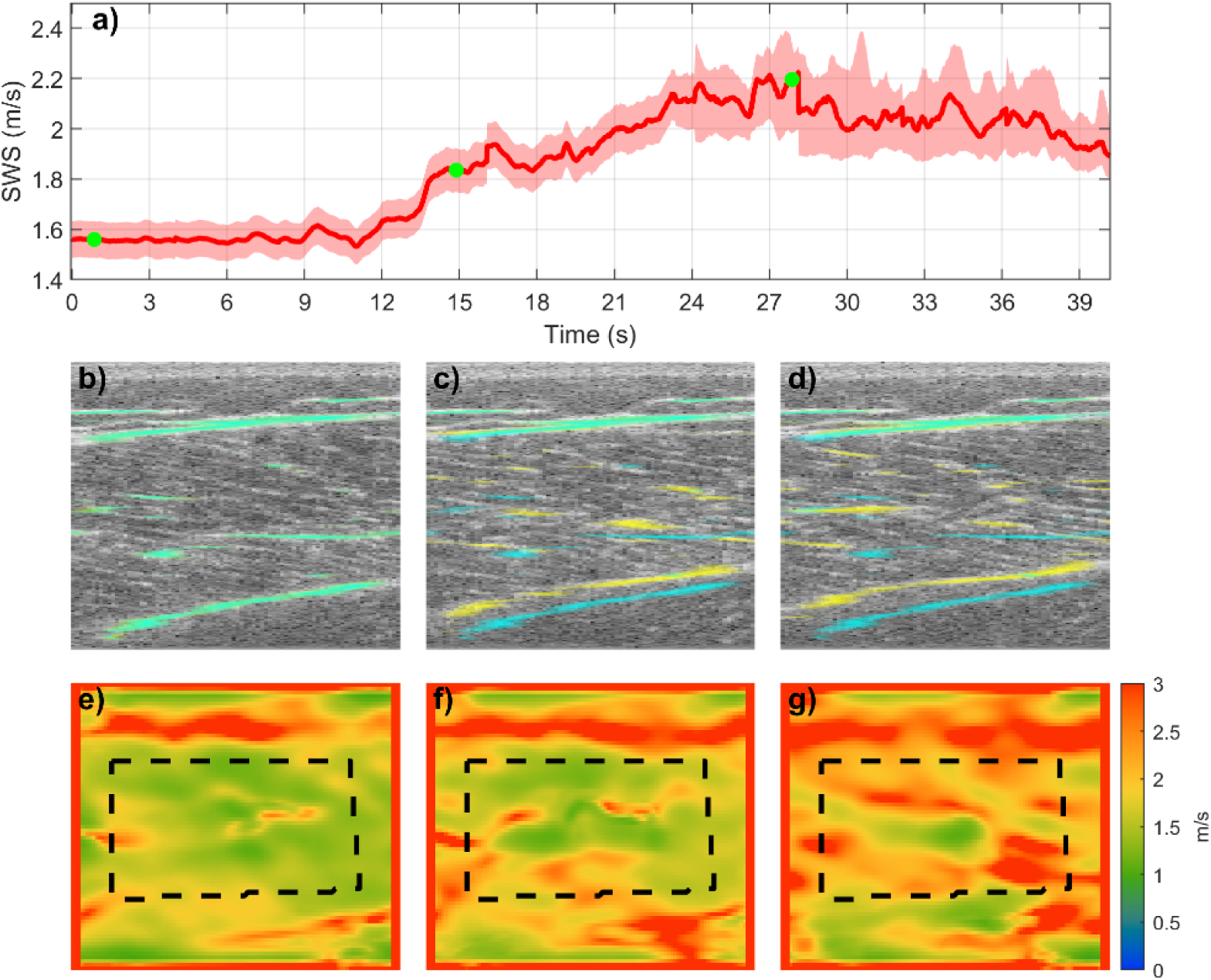
Temporal and spatial response of VL muscle to TVR stimulation in a representative volunteer. (a) Mean SWS time course in the selected ROI during 40 s of TVR stimulation at moderate amplitude (400 µm) with the shaded area indicating the standard deviation. (b–d) B-mode ultrasound images at 0.5 s (baseline), 15 s (around beginning of contraction), and 27 s (sustained contraction). Muscle boundaries and prominent fibers at each time point are marked in yellow, while those from the baseline (0.5LJs, panel b) are superimposed in semi-transparent blue on panels b–d. Areas where yellow and blue overlap appear green; movement relative to baseline is revealed by the non-overlapping regions in the original colors. (e–g) Corresponding spatially resolved SWS maps at 0.5, 15, and 27 s. Increased stiffness (higher SWS) within the ROI, outlined by the dashed line, correlates with muscle contraction visualized in B-mode.

Corresponding B-mode images at 0.5, 15, and 27 s are shown in panels (b–d). Muscle boundaries and most prominent fibers are marked in yellow. The t=0.5 s image (Figure 3b) is superimposed in transparent blue on Figures 3b–3d. Overlapping yellow and blue appear green; movement reveals original colors. At 0.5 s (Figure 3b), the VL muscle showed a resting morphology. Slight structural changes appeared at 15 s (Figure 3c) and became more pronounced at 27 s (Figure 3d), indicating muscle contraction by a decreased overall muscle thickness, fiber shortening and fiber thickening, resulting in an altered echogenicity under sustained reflex activation.

Figures 3e–3g display SWS maps for 0.5, 15, and 27 s. At 0.5 s (Figure 3e), SWS was uniformly low within the ROI, reflecting relaxed tissue. At 15 s (Figure 3f), stiffness increased with initial muscle activation, reaching significantly higher values at 27 s (Figure 3g).

### 3.2. SWS Across Conditions

At baseline (rest), mean VL SWS was 1.66 + 0.11□m/s, with a 10th percentile (Q10) of 1.57□m/s and a 90th percentile (Q90) of 1.76□m/s. SWS increased with muscle activation due to VC or TVR (Table 1).

**Table 1:** SWS summary across conditions. IDR = interdecile range (Q10 – Q90)

| Condition | IDR<br>(m/s) | Mean ± SD<br>(m/s) | $\Delta$ from rest<br>(m/s) | Increase<br>% | $p$-value vs<br>rest | Cohen's<br>d |
| --- | --- | --- | --- | --- | --- | --- |
| **Base line (Rest)** | 1.57 – 1.76 | 1.66 ± 0.11 | — | — | — | — |
| **VC 15 N** | 1.63 – 1.89 | 1.79 ± 0.12 | +0.13 | 8 | $< 10^{-5}$ | 1.11 |
| **VC 22 N** | 1.84 – 2.05 | 1.93 ± 0.13 | +0.27 | 16 | $< 10^{-13}$ | 2.25 |
| **VC 37 N** | 2.10 – 2.24 | 2.16 ± 0.12 | +0.49 | 30 | $< 10^{-16}$ | 4.43 |
| **TVR 200 $\mu\text{m}$** | 1.84 – 2.00 | 1.93 ± 0.13 | +0.27 | 17 | $< 10^{-9}$ | 2.27 |
| **TVR 400 $\mu\text{m}$** | 2.05 – 2.28 | 2.18 ± 0.14 | +0.52 | 31 | $< 10^{-15}$ | 4.25 |

A repeated-measures ANOVA revealed a highly significant effect of condition on SWS (F(5,120) = 99.12, *p*< 0.001). Post-hoc *t*-tests and Wilcoxon signed-rank tests confirmed all VC and TVR conditions exceeding baseline SWS values (*p* < 0.001 for each comparison). However, VC at 22 N versus TVR with 200□µm amplitude and VC at 37 N versus TVR with 400□µm did not differ significantly, indicating that voluntary and vibration-induced contractions elicited equivalent stiffness changes at moderate and high intensities. Table 1 summarizes SWS distribution, effect sizes, and *p*-values relative to baseline.

Figure 4 displays box plots of SWS measurements across conditions. Individual subject data are connected by lines, clearly illustrating the within-subject trend of increasing stiffness from the rest condition to VC and TVR.

**Figure 4:**
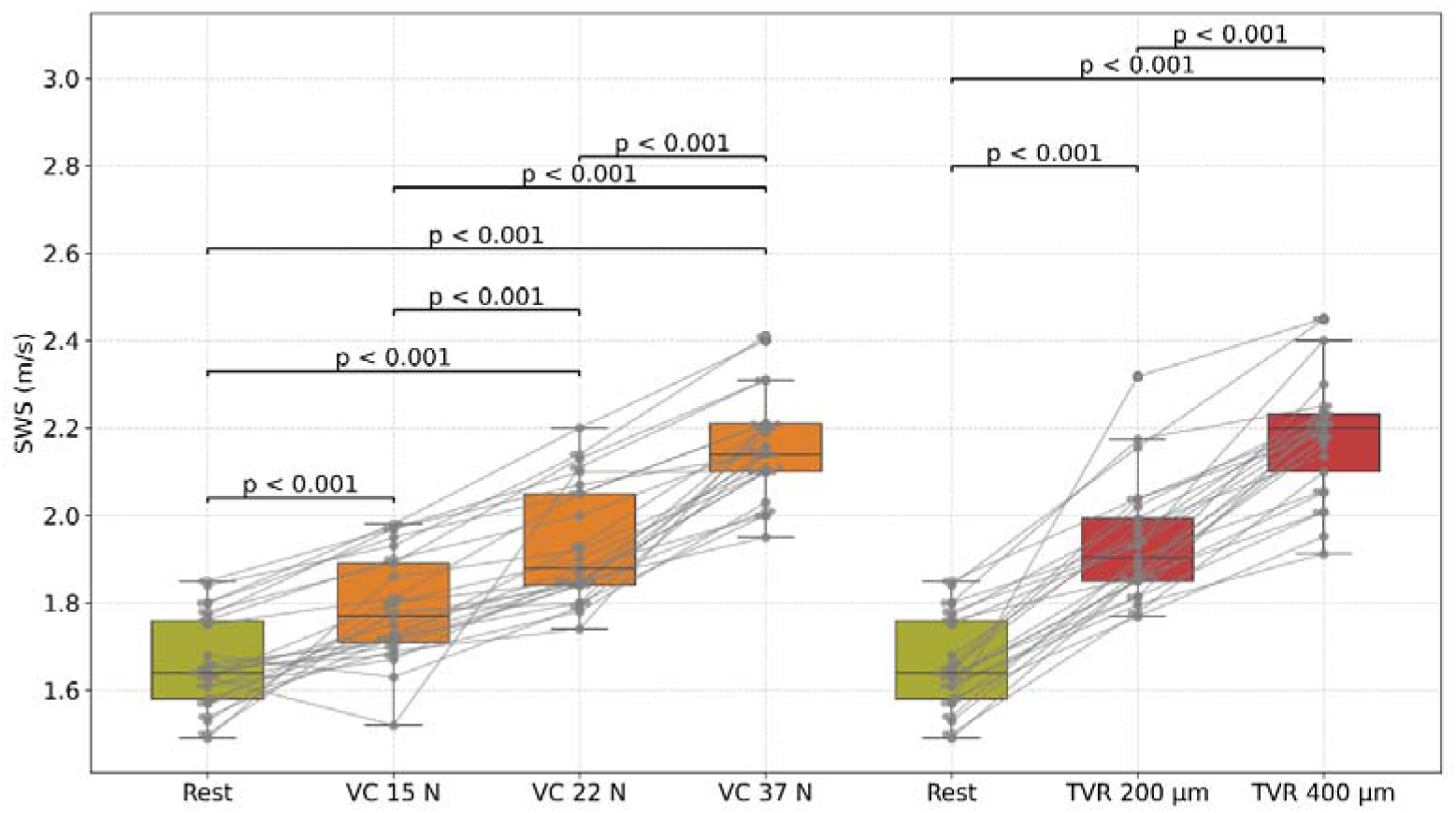
Group statistical plots of SWS values across six conditions (Rest, VC 15 N, VC 22 N, VC 37 N, TVR 200□µm, TVR 400□µm). Connecting lines represent individual subject trajectories. Note the similar values between VC 22 N and TVR 200□µm, as well as between VC 37 N and TVR 400□µm.

### 3.3. Correlation Analysis Between VC and TVR

We next examined whether higher VC SWS values correlated with higher TVR SWS. Indeed, a significant correlation was observed for the high-intensity condition (VC 37 N versus TVR 400□µm, r = 0.67, *p* < 0.001), indicating that participants with higher SWS under VC also tended to exhibit higher SWS under TVR. This relationship is illustrated in Figures 4 and 5a. In contrast, under moderate-intensity conditions (VC at 22 N versus TVR at 200□µm), no correlation (r > 0.5) was found between the moderate-level intensities (r = 0.30, *p* = 0.145), suggesting that the stiffening effect of moderate VC and TVR may vary across individuals (Figure 5b).

**Figure 5:**
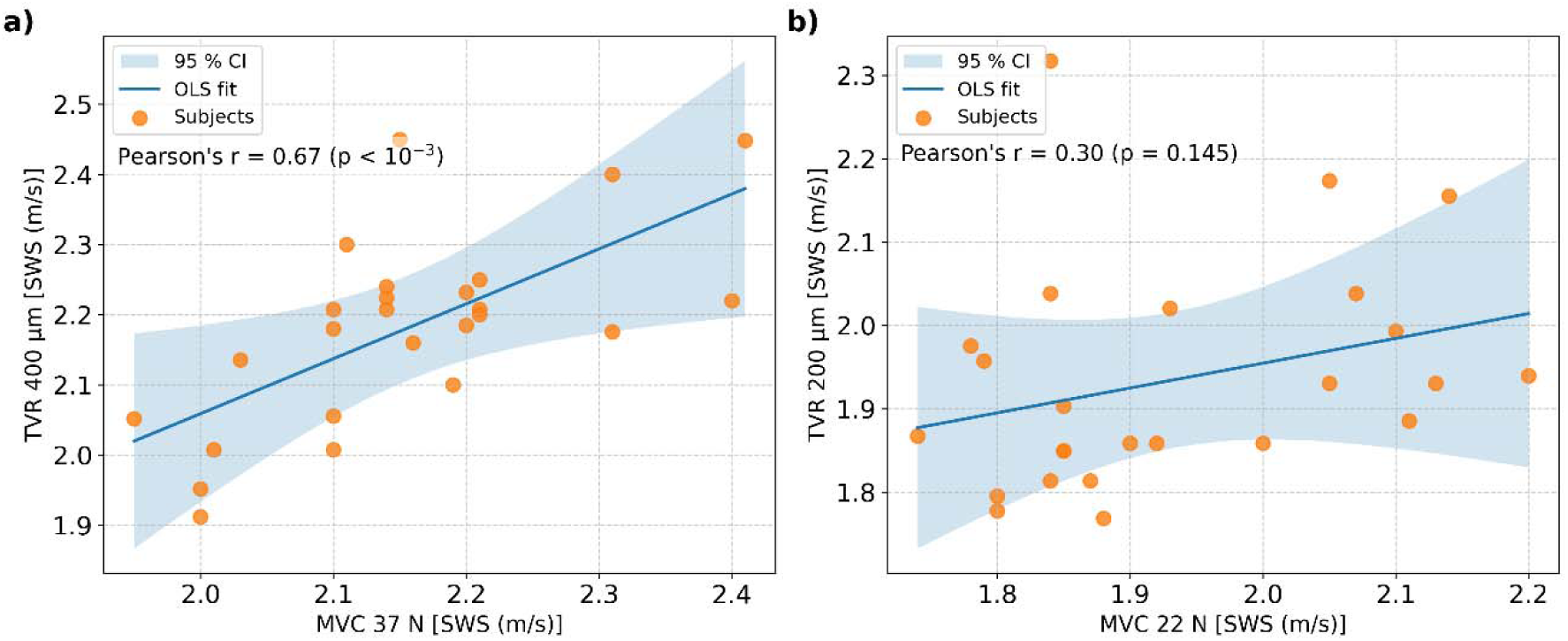
Scatter plots of SWS measurements during TVR and VC conditions. (a) VC 37□N versus TVR 400□µm; (b) VC 22□N versus TVR 200□µm. A clear correlation is observed in panel (a). The blue line represents the ordinary least squares (OLS) fit, with the shaded area indicating the 95% confidence interval (CI).

### 3.4. Subgroup Analyses

To assess the influence of demographic and physiological factors on stiffness, subgroup analyses were conducted for BMI, sex, age, and leg dominance. BMI analysis showed underweight participants had lower baseline SWS but similar or greater increases with high force/vibration, while overweight participants had higher baseline SWS values but similar SWS changes. No significant sex differences were found in baseline or activated SWS, suggesting that the increase in stiffness was dependent on contraction intensity. Older adults exhibited slightly higher baseline stiffness but achieved comparable SWS under peak loads. Leg dominance showed no difference in baseline or activated SWS, indicating similar responses to identical loads or vibration. Overall, no significant differences (p>0.05) were found across these subgroups for either baseline SWS or magnitude of stiffness changes. However, these findings are limited by small subgroup sizes. Table 3 provides a summary of these comparisons for rest and peak activations at VC (37 N) or TVR (400 µm).

**Table 3:** Subgroup Comparisons: Baseline vs. High-Intensity SWS.

| Factor | Group | Rest (m/s) | VC37 or TVR 400 μm (m/s) | *p*-value |
| --- | --- | --- | --- | --- |
| BMI | Underweight <18.5 (N=2) | 1.61 ± 0.06 | 2.23 ± 0.03 | —<sup>a</sup> |
| | Normal 18.5-25 (N=19) | 1.66 ± 0.09 | 2.15 ± 0.11 | 0.28 |
| | Overweight <25 (N=4) | 1.71 ± 0.07 | 2.12 ± 0.12 | 0.30 |
| Sex | Male (N=17) | 1.65 ± 0.10 | 2.17 ± 0.11 | 0.36 |
| | Female (N=8) | 1.68 ± 0.12 | 2.13 ± 0.15 | 0.40 |
| Age | Younger < 30 (N=14) | 1.65 ± 0.10 | 2.15 ± 0.14 | 0.41 |
| | Middle 30–45 (N=7) | 1.67 ± 0.10 | 2.19 ± 0.10 | 0.32 |
| | Older > 45 (N=4) | 1.70 ± 0.07 | 2.11 ± 0.09 | 0.24 |
| Leg Dominance | Dominant (N=20) | 1.66 ± 0.11 | 2.15 ± 0.12 | 0.48 |
| | Non-Dominant (N=5) | 1.68 ± 0.11 | 2.19 ± 0.10 | 0.58 |

<sup>a</sup>Too few underweight subjects (N=2) for meaningful statistical testing.

## 4. Discussions

This study demonstrates effectiveness of TVR for VL muscle function assessment by THE. To the best of our knowledge, our study showed for the first time that TVR increases skeletal muscle stiffness similarly to voluntary contraction. TVR with 400 µm amplitude caused a similar ∼30% increase of muscle stiffness than VC at 37 N. This supports our hypothesis that TVR is a reliable, objective muscle activation method, overcoming the subjectivity of voluntary efforts.

TVR is known to be mediated by muscle spindle Ia afferent activation, causing involuntary motor unit recruitment (Romaiguère et al., 1993). While prior work addressed TVR variability based on stimulus parameters (Eklund & Hagbarth, 1966) and explored neuromechanical mechanisms like muscle waves (Guang et al., 2018) and frequency-dependent synchronization (Martin & Park, 1997), our study directly quantified resulting mechanical property changes using non-invasive THE. We selected 100 Hz TVR frequency because, first, this stimulus has been known to elicit muscle activation (Eklund & Hagbarth, 1966; Poenaru et al., 2016) and, second, this frequency could be well synchronized to the frame rate of our THE scanner. Our observation that higher vibration amplitude increases stiffness aligns with the known TVR dose-response (Eklund & Hagbarth, 1966).

THE allowed time-resolved SWS mapping across the VL, revealing reflex activation dynamics. Figure 3 shows progressive SWS increase over seconds during TVR, correlating with visible morphological changes (fiber shortening, tissue thinning) on B-mode imaging. This dynamic-spatial SWS capture is an advantage of THE over conventional elastography methods as recently demonstrated for the assessment of skeletal muscle stiffness during prolonged contractions and recovery (Yang et al., 2024).

A moderate positive correlation between stiffness during VC (37 N) and TVR (400 µm) suggests a link between voluntary activation capacity and reflex responsiveness at higher intensities. However, the lacking sensitivity of THE at lower intensities warrants further investigation of factors influencing voluntary and reflex activation of skeletal muscle.

Our subgroup analyses found no relevant influence of sex, age, BMI, or leg dominance on SWS to VC or TVR in our asymptomatic cohort. This contrasts with some studies reporting age/sex effects on muscle stiffness using other methods (Eby et al., 2015; Wang et al., 2017; Chodock et al., 2020). However, limited subgroup sample sizes, especially for age/BMI extremes, prevent definitive conclusions here. Moreover, THE measures stiffness in a lower dynamic range than ARFI-based methods (Tzschätzsch et al., 2018, Burkhardt et al., 2020) preventing a direct comparison of our results with these studies.

Clinically, TVR-THE’s ability to induce and measure muscle stiffness is promising for objective muscle function evaluation in individuals unable to perform reliable VC due to pain, neurological impairment (e.g., stroke, spasticity, Parkinson’s disease as studied by Burke et al., 1972; McPherson et al., 2018), or lack of cooperation. While local vibration has therapeutic applications (Poenaru et al., 2016), its combination with elastography for diagnostic/monitoring is novel. Integrating TVR could standardize muscle activation for better assessment comparability and potentially track rehabilitation or disease status, similar to elastography in inflammatory myopathies (Alfuraih et al., 2019). Furthermore, the technical setup of TVR is leaner than that of VC stimulation as no force measurement device or normalized counter-force has to be involved. A small additional shaker can be easily integrated into clinical sonography protocols.

Several limitations exist. This study focused on a co-activated VL muscle in asymptomatic participants. Our findings may differ in other muscles or clinical populations. Using a single TVR frequency is a limitation. A range of frequencies might elicit a more robust activation reflex, which however is constrained by the THE sampling rate. Preliminary tests with 200 Hz TVR in a subgroup of volunteers were not successful at this stage. VC was based on fixed external loads, not percentage of maximal voluntary contraction. The absolute VC levels might have added variability related to individual muscle strength and state of training. However, since our feasibility study included only asymptomatic and relatively young subjects, we do not expect this to change our conclusions. Anisotropy was also ignored in this study. However, since muscle is an anisotropic medium (Ngo et al., 2022), and SWS differs with fiber direction, accounting for anisotropy could lead to more consistent results. Finally, the modest sample size of 25 asymptomatic volunteers limits in-depths subgroup analyses and the generalizability to clinical populations like stroke or spasticity patients, where TVR responses may differ.

## 5. Conclusions

This study demonstrated that THE effectively quantifies VL stiffness changes induced by both VC and TVR. Notably, TVR stimulation at 100 Hz with moderate amplitude (400 µm) resulted in stiffness increases similar to VC in response to 37 N load, indicating TVR’s potential to substitute more subjective voluntary methods in functional muscle elastography. Our findings suggest TVR-THE as an objective method for assessing muscle activation and monitoring muscular disease treatment based on stiffness changes. THE may offer significant clinical value, particularly for evaluating muscle dysfunction in individuals unable to perform reliable VC exercises. Further research is needed to explore TVR-THE across different muscles and clinical populations.

## Data Availability

All data produced in the present study are available upon reasonable request to the authors

## References

Brandenburg, J. E., Eby, S. F., Song, P., Zhao, H., Brault, J. S., Chen, S., & An, K. N. (2014). Ultrasound elastography: the new frontier in direct measurement of muscle stiffness. Archives of Physical Medicine and Rehabilitation, 95(11), 2207–2219.

Burke, D., Andrews, C. J., & Lance, J. W. (1972). Tonic vibration reflex in spasticity, Parkinson’s disease, and normal subjects. Journal of Neurology, Neurosurgery & Psychiatry, 35(4), 477–486.

Burkhardt, C., Tzschaetzsch, H., Schmuck, R., Bahra, M., Juergensen, C., Pelzer, U., … & Garcia, S. R. M. (2020). Ultrasound time-harmonic elastography of the pancreas: reference values and clinical feasibility. Investigative Radiology, 55(5), 270–276.

Catheline, S., Gennisson, J. L., Delon, G., Fink, M., Sinkus, R., Abouelkaram, S., & Culioli, J. (2004). Measurement of viscoelastic properties of homogeneous soft solid using transient elastography: An inverse problem approach. The Journal of the Acoustical Society of America, 116(6), 3734–3741.

Cohen J. Statistical Power Analysis for the Behavioural Sciences. Lawrence Earlbaum Associates: Hillside, NJ, USA, 1988; pp. 77–83, 278-280.

Deffieux, T., Gennisson, J. L., Tanter, M., & Fink, M. (2008). Assessment of the mechanical properties of the musculoskeletal system using 2-D and 3-D very high frame rate ultrasound. IEEE transactions on ultrasonics, ferroelectrics, and frequency control, 55(10), 2177–2190.

Doyley, M. M., & Parker, K. J. (2014). Elastography: general principles and clincial applications. Ultrasound clinics, 9(1), 1.

Eby, S. F., Song, P., Chen, S., Chen, Q., Greenleaf, J. F., & An, K. N. (2013). Validation of shear wave elastography in skeletal muscle. Journal of Biomechanics, 46(14), 2381–2387.

Eklund, G., & Hagbarth, K. E. (1966). Normal variability of tonic vibration reflexes in man. Experimental Neurology, 16(1), 80–92.

Fung, Y. C. (2013). Biomechanics: mechanical properties of living tissues. Springer Science & Business Media.

Gennisson, J. L., Deffieux, T., Macé, E., Montaldo, G., Fink, M., & Tanter, M. (2010). Viscoelastic and anisotropic mechanical properties of in vivo muscle tissue assessed by supersonic shear imaging. Ultrasound in medicine & biology, 36(5), 789–801.

Guang, H., Ji, L., & Shi, Y. (2018). Focal vibration stretches muscle fibers by producing muscle waves. IEEE Transactions on Neural Systems and Rehabilitation Engineering, 26(4), 839–846.

Hall, T. J., Milkowski, A., Garra, B., Carson, P., Palmeri, M., Nightingale, K., … & Zhao, H. (2013, July). RSNA/QIBA: Shear wave speed as a biomarker for liver fibrosis staging. In 2013 IEEE International Ultrasonics Symposium (IUS) (pp. 397–400). IEEE.

Kasai, C., Namekawa, K., Koyano, A., & Omoto, R. (1985). Real-time two-dimensional blood flow imaging using an autocorrelation technique. IEEE Transactions on Sonics and Ultrasonics, 32(3), 458–464.

Manduca, A., Lake, D. S., Kruse, S. A., & Ehman, R. L. (2003). Spatio-temporal directional filtering for improved inversion of MR elastography images. Medical image analysis, 7(4), 465–473.

Marín, P. J., & Rhea, M. R. (2010). Effects of vibration training on muscle power: A meta-analysis. Journal of Strength and Conditioning Research, 24(3), 871–878.

Martin, B. J., & Park, H. S. (1997). Analysis of the tonic vibration reflex: influence of vibration variables on motor-unit synchronization and fatigue. European Journal of Applied Physiology and Occupational Physiology, 75(6), 504–511.

Meyer, T., Wellge, B., Barzen, G., Knebel, F., Hahn, K., Elgeti, T., … & Sack, I. (2024). Point-of-care cardiac elastography with external vibration for quantification of diastolic myocardial stiffness. medRxiv, 2024-01.

Ngo, H. H. P., Poulard, T., Brum, J., & Gennisson, J. L. (2022). Anisotropy in ultrasound shear wave elastography: an add-on to muscles characterization. Frontiers in Physiology, 13, 1000612.

Parker, K. J., Doyley, M. M., & Rubens, D. J. (2010). Imaging the elastic properties of tissue: the 20 year perspective. Physics in medicine & biology, 56(1), R1.

Poenaru, D., Cinteza, D., Petrusca, I., Cioc, L., & Dumitrascu, D. (2016). Local application of vibration in motor rehabilitation–scientific and practical considerations. Maedica, 11(3), 227.

Sack, I. (2023). Magnetic resonance elastography from fundamental soft-tissue mechanics to diagnostic imaging. Nature Reviews Physics, 5(1), 25–42.

Sawilowsky, S. S. (2009). New effect size rules of thumb. Journal of modern applied statistical methods, 8(2), 26.

Song, P., Zhao, H., Manduca, A., Chen, S., & Greenleaf, J. F. (2013). Comb-push ultrasound shear elastography (CUSE) with various ultrasound push beams. IEEE Transactions on Medical Imaging, 32(8), 1435–1447.

Tang, C., Gao, Y., Chen, J., Liu, C., & Li, Z. (2020). Ultrasound shear wave elastography and its association with muscle force and muscle activity in vivo. Journal of Healthcare Engineering, 2020, 8853573.

Tzschätzsch, H., Guo, J., Dittmann, F., Hirsch, S., Barnhill, E., Jöhrens, K., & Sack, I. (2016b). Two-dimensional time-harmonic elastography of the human liver and spleen. Ultrasound in Medicine & Biology, 42(11), 2562–2571.

Tzschätzsch, H., Guo, J., Hirsch, S., Hopfner, F., Schrader, F., Guo, J., Dittmann, F., et al. (2016a). Tomoelastography by multifrequency wave number recovery for in vivo detection of focal lesions. Medical Image Analysis, 30, 1–10.

Tzschätzsch, H., Kreft, B., Schrank, F., Bergs, J., Braun, J., & Sack, I. (2018). In vivo time-harmonic ultrasound elastography of the human brain detects acute cerebral stiffness changes induced by intracranial pressure variations. Scientific reports, 8(1), 17888.

Tzschätzsch, H., Chandia, S. K., & Meyer, T. (2024). Methods and approaches in ultrasound elastography. In Quantification of biophysical parameters in medical imaging (pp. 323–344). Cham: Springer International Publishing.

Xu, J., Hug, F., & Fu, S. N. (2018). Stiffness of individual quadriceps muscle assessed using ultrasound shear wave elastography during passive stretching. Journal of sport and health science, 7(2), 245–249.

Yang, Y., Shahryari, M., Meyer, T., Garcia, S. R. M., Görner, S., Majd, M. S., … & Tzschätzsch, H. (2024). Explorative study using ultrasound time-harmonic elastography for stiffness-based quantification of skeletal muscle function. Zeitschrift für Medizinische Physik, in press (DOI: 10.1016/j.zemedi.2024.03.001)

Zaidell, L. N., et al. (2013). Experimental evidence of the tonic vibration reflex during whole-body vibration of the loaded and unloaded leg. PLoS One, 8(12), e85090.

